# Building a culture of developing annual health work plan based on data: Critical health system related factors influencing data utilization in Bukedi region, Uganda

**DOI:** 10.1101/2025.03.31.25324989

**Authors:** Job Cherukut, Swaibu Zziwa, Naziru Rashid, Aremu Abudulmujeeb Babatunde, Hussein Mukasa Kafeero, Abdul Walusansa, Ziada Nankinga, Sumayah Nakitende, Kharim Mwebaza Muluya

## Abstract

Utilization of routine health data for Annual Health Work Plan (AHWP) development is crucial for effective health planning. However, data use remains a challenge in many health facilities, especially in low-resource settings like the Bukedi region of Uganda. In order to assess data utilization for AHWP development and identify health system related factors influencing data use among healthcare workers in Bukedi region, a cross-sectional study was conducted and 313 healthcare workers in Bukedi region health centres were interviewed. Data on data utilization practices and health system related factors were collected and analyzed using STATA *version 14*. Multivariate analysis identified significant health system related factors influencing data utilization for AHWP development. Most respondents were female (59.74%), married (77.96%), Catholic (46.65%), residing outside health centre quarters (82.43%) and health workers without medical records qualification were 91.69%. The level of data utilization for AHWP development was low at 31%. Multivariate analysis showed significant health system-related factors and it included regular facility meetings (AOR: 0.4; 95% CI: 0.23 – 0.92; P=0.028), poor data quality (AOR: 0.0003; 95% CI: 0.00003 – 0.004; P=0.000), feedback from the Ministry of Health (AOR: 47.4; 95% CI: 19.5 – 115.02; P=0.000), leadership culture (AOR: 42.2; 95% CI: 20.6 – 86.6; P=0.000), availability of trained staff (AOR: 300.1; 95% CI: 98.9 – 911.2; P=0.000), and internet availability (AOR: 0.07; 95% CI: 0.009 – 0.49; P=0.008). Data utilization for AHWP development in Bukedi health centres remains low and is influenced by various health system related factors, such as lack of skills, inadequate training, insufficient leadership support, poor data quality, and infrastructure gaps. Addressing these barriers through training, leadership engagement, and infrastructure improvements is crucial to enhance data-driven health planning.

## Introduction

Globally, the utilization of routine health data is fundamental to improving health system efficiency and achieving universal health coverage (UHC). Health information systems (HIS) provide a foundation for data-driven decision-making, which enables health facilities to identify health priorities, allocate resources effectively, and monitor progress on key health indicators. The World Health Organization (WHO) has emphasized data use in health systems as a cornerstone of the Health System Strengthening Framework, aligning with Sustainable Development Goal (SDG) 3, which aims to ensure healthy lives and promote well-being for all ages [1]. In high-income countries, the integration of HIS has allowed health facilities to make evidence-based decisions, resulting in improved health outcomes and more efficient resource allocation [2]. For example, Ireland’s Health Information and Quality Authority (HIQA) reports a 30% increase in the use of data for health planning due to structured data governance frameworks, which enhance data quality and accessibility for decision-makers.

Across continents, countries have made notable strides in data-driven health planning by enhancing HIS infrastructure. Japan and Singapore, for instance, have implemented advanced health information systems that prioritize data utilization for health planning, resource allocation, and policy development [3]. Singapore’s Ministry of Health uses HIS to perform comprehensive data analyses that shape national health strategies and support the efficient delivery of healthcare services. The intercontinental disparities in HIS functionality, however, are evident in the challenges faced by low- and middle-income countries (LMICs) in implementing similar systems. Many LMICs still struggle with limited technical resources, low data literacy, and insufficient infrastructure, all of which hinder the effective use of HIS for planning and decision-making [4]. In response, countries like Ethiopia and Nigeria have included HIS capacity building in their national health agendas, though implementation remains constrained by these systemic challenges [5].

In sub-Saharan Africa, HIS adoption and data utilization have been prioritized, but significant gaps persist in integrating data into health planning. Despite ongoing initiatives to enhance data literacy and infrastructure, HIS effectiveness remains limited due to factors such as inconsistent data quality, inadequate training, and infrastructural constraints. The Ethiopian Health Sector Transformation Plan (HSTP) highlights HIS as a critical component for health system improvement but notes persistent barriers, including insufficient data management skills and inadequate feedback mechanisms for health workers [6]. In Nigeria, the National Health Plan includes data utilization targets, yet challenges in infrastructure, technology, and data quality limit the impact of HIS on health planning [4]. These limitations have contributed to suboptimal health outcomes, particularly in rural and underserved regions where resource allocation and healthcare delivery critically depend on effective data use.

In East Africa, similar challenges affect data utilization in health planning, though recent efforts have aimed to enhance health system responsiveness through better data practices. Studies in Kenya show that healthcare facilities with trained staff, regular feedback mechanisms, and supportive leadership, report higher rates of data use for health planning [7]. In Tanzania, data utilization in health facilities has been supported by donor-funded programs that emphasize data quality improvement and health worker training. However, healthcare workers in these settings often view data entry as an administrative task, limiting the role of data in informing strategic decisions. Moreover, the region’s reliance on donor support for HIS initiatives raises concerns about the sustainability of data-driven practices in health planning. As in other LMICs, the development of HIS infrastructure and consistent training remains essential for realizing the full potential of data utilization in health system strengthening [4].

In Uganda, routine health data utilization for planning remains limited, particularly in rural regions like Bukedi, where healthcare facilities face significant health challenges. The Ugandan Ministry of Health (MoH) has implemented HIS across health facilities and introduced tools such as the HMIS001 tool and Primary Health Care (PHC) grant guidelines, which mandate data use for identifying poor-performing indicators in Annual Health Work Plans (AHWPs) [8]. Despite these efforts, health indicators such as antenatal care (ANC), postnatal care (PNC), and family planning (FP) coverage remain below national targets, reflecting the limited impact of data-driven planning in regions like Bukedi region [9]. Low performance of Bukedi region (54.2%) in health indicators has relegated the region to the tail end of the national rankings, and below the 68% national average score [10].

The limited capacity for data utilization in Uganda, particularly in rural districts, highlights a critical gap in data-driven health planning. Importantly, health facility data on performance indicators remain available, though little is known if facilities utilize it in guiding key decisions during AHWP development. It is not clear why health facilities struggle to meet the required data utilization rates, contributing to poor performance and ineffective health interventions often leading to missed opportunities for quality improvement and lack of preparedness for emerging health issues in health centres among others [11]. This study aimed at assessing the health system related factors influencing data utilization for AHWP development in Bukedi region, with a focus on identifying actionable strategies to strengthen health planning and service delivery in the region.

## Materials and methods

### Study setting

The study was conducted in Butebo and Pallisa districts found in eastern Uganda, which are part of the seven districts that form Bukedi region. Butebo District formally curved out of Pallisa district in 2017 is bordered to the north by Ngora, Kumi and Bukedea Districts, Mbale District to the east, Budaka District to the south and Pallisa District to the west while Pallisa District is bordered by Serere District in the Northwest, Ngora in the North, Kumi in the Northeast, Bukedea, Butebo and Budaka Districts in the east, Kibuku and Kaliro in the south.

They have a number of health centres at the different levels, including government-run facilities, private clinics, and private not for profit organizations totaling to 40 facilities as per Ministry of Health reporting system. Furthermore, these districts share similar features and are predominantly rural, with a population that relies heavily on the health centres for their healthcare needs. They have several sub-counties, with nearly all having its own health centres serving the surrounding communities. Butebo and Pallisa districts face a high burden of preventable diseases such as malaria, diarrhea, urinary tract infections, HIV/AIDS, and tuberculosis among others that has strained the health budget available. Similarly, their general performance in the different health indicators on the maternal child health and nutritional cascade such as ANC1 first trimester and ANC4 visits, PNC, FP and child immunization were very poor and puts the two districts at the tail end.

### Study population

Health workers working in selected health centres in Bukedi region which were receiving PHC grants guidelines and indicative figures to develop AHWPs were considered. The health workers in health centres were key in responding to the structured questions.

### Eligibility criteria

Only health workers working in health centres in Butebo and Pallisa district that were receiving PHC grants who were supposed to develop AHWPs and met the sampling criteria were considered. Staff who were working in these health centres for at least one year and were willing to provide informed consent were the respondents of the study. Meanwhile health staff who were unwilling to provide informed consent, severely sick, and those mentally unstable during the study period, were excluded from this study.

### Study design

The study utilized a cross-sectional design which employed quantitative data collection method to assess data utilization for AHWP development and the influencing health system related factors in health centres in the selected districts of Butebo and Pallisa. This design allowed for the collection of data at a single point in time from a representative sample of the study population.

### Sample size estimation

A total of 326 health workers from 28 health centres were randomly selected from the two districts in Bukedi region. The selection of respondents from health centres was based on the modified Kish formula for a single proportion, adapted for finite populations.

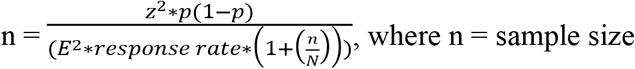

Z = Z-score (1.96 for 95% confidence which the study shall consider)

p = estimated proportion of data utilization for AHWPs (unknown so we took 50%) E = desired level of precision (margin of error) which is 5% or 0.005

Response rate = proportion of participants who respond to the study and provide usable data which is 0.6 or 60% that this study shall consider.

N = total size of the finite population (population size) which is 491

n/N = sampling fraction (the ratio of the sample size to the population size) The values are then plugged in as; n = 326

So, the estimated sample size is approximately 326

This calculation was used because it assumes a finite population correction, which is useful when the population size is known. Furthermore, the sample size was adjusted for each facility level using the formula as shown below;

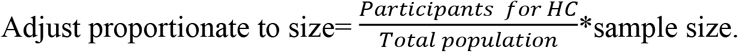

The sample was adjusted for each facility level as shown in the Table 1.

**Table 1:**
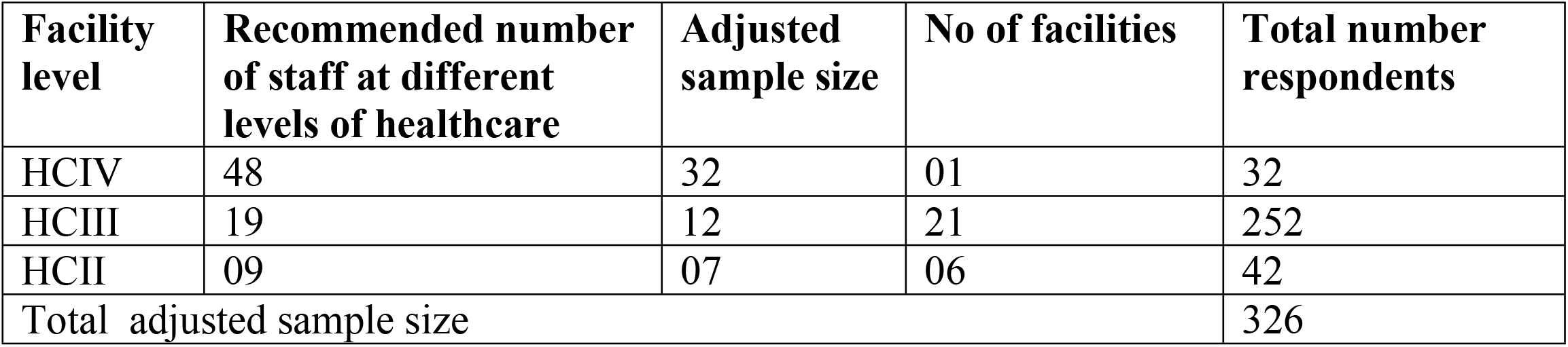
Adjusted sample size of health centres.

| Facility level | Recommended number of staff at different levels of healthcare | Adjusted sample size | No of facilities | Total number respondents |
| --- | --- | --- | --- | --- |
| HCIV | 48 | 32 | 01 | 32 |
| HCIII | 19 | 12 | 21 | 252 |
| HCII | 09 | 07 | 06 | 42 |
| Total adjusted sample size |  |  |  | 326 |

### Sampling technique / procedure

Stratified sampling of health centres (HCs) into three levels of HCIV, HCIII, and HCIIs was adopted. One HCIV was purposively selected, however, the 21 HCIIIs and 06 HCIIs were randomly selected from the 28 and 11 health centres respectively using simple random sampling.

This ensured a representative sample of the population in consideration of the resources and time available for data collection. Stratified sampling minimized bias and ensured that the results are generalizable to the population of health centres in the districts of Butebo and Pallisa.

For the study respondents from these participating health centres, simple random sampling technique was used to select participants for the study by listing the names of prospective participants in each health centre. A random number was generated from a set of random numbers within the range of the total number of participants which was used to select the names corresponding to the generated random numbers. This was repeated for all other health centres that were included in the study. Simple random sampling guaranteed every staff member had an equal chance of being selected, making the sample a representative of the population and it reduced sampling bias and errors ensuring that the results accurately reflect the population of health centre staff.

### Data collection methods and tools

The data collection method was administration of structured questionnaires. The questionnaires were administered to study respondents by the researcher and trained research assistants. The whole exercise of data collection started on 7^th^ July, 2024 and ended on 1^st^ August, 2024 after the IRB clearance.

A structured questionnaire was used to collect data. A pretested tool with relevant questions was adopted and modified from the MEASURE Evaluation Manual: Tools for Data Demand and Use in the Health Sector (Version 2) and further developed based on Ministry of Health planning tool HMIS001 and PHC guidelines for relevant data utilization parameters that inform annual work planning in health facilities. This tool was used to guide data collection for all variables. The research instrument was adjusted based on pretesting results to ensure that data collection was systematic, comprehensive, and rigorous, thereby maintaining the quality and integrity of the research findings.

### Study variables and their measurements

The dependent variable in this study was the utilization of data for developing Annual Health Work Plans (AHWPs). It was measured by reviewing participants’ self-reported responses on the use of available health data to support decision-making and activity planning for AHWPs. A respondent was classified as having utilized data for annual health work planning if they achieved an average score of 80% or above based on specific key assessment criteria. These criteria were aligned with the health sub-program grant, budget guidelines, and local government implementation protocols.

The independent variable in this study is health system related factors. Health system related factors were measured using binary responses (Yes/No, 1/0), including departmental staff meetings (1 = Present, 0 = Absent), feedback on facility performance based on data (1 = Received, 0 = Not received), leadership promoting data use (1 = Present, 0 = Absent), adequate trained staff (1 = Available, 0 = Not available), data quality challenges (1 = Present, 0 = Absent), availability of standard guidelines for AHWPs (1 = Available, 0 = Not available) and internet availability (1 = Available, 0 = Not available).

### Quality control measures

To ensure the quality of the data collected, several quality control measures were implemented. The questionnaires were reviewed and reformatted based on the pretest result. Research assistants were trained on data collection tools and techniques to ensure familiarity with the tools. Supervision and monitoring of data collection activities was done regularly.

### Validity

Validity in this study was ensured through content, face, and criterion validity. *Content validity* was achieved by designing data collection tools that covered all parameters and indicators of data utilization, based on the modified Ministry of Health tool HMIS001 and PHC guidelines, with expert consultation to confirm comprehensiveness and relevance. While as *face validity* was established by ensuring that the tools appeared to measure the intended variables and were aligned with the research concepts for accuracy. Lastly, *criterion validity* was ensured by verifying theoretical alignment, ensuring that the tools captured the underlying theoretical frameworks, and achieving a high response rate of 95.7%, which provided strong validity to the findings. According to Creswell and Creswell, a response rate of 90% or higher is considered excellent, while a rate of 70–89% is considered good [12]. Therefore, the response rate in this study was excellent, further strengthening the validity of the findings.

### Reliability

Based on Amin’s recommended value of 0.70, the Cronbach’s Alpha (α) coefficient indicated that the instrument was reliable [13]. Pilot testing of the data collection tools was conducted with 10% of the sample size to ensure that the questions and measures were clear. The confirmed results remained consistent over time, ensuring temporal stability. This testing was carried out in one Health Centre IV in the Bukedi region.

A pretest was conducted using Cronbach’s Alpha to assess the validity of the instruments. This initial phase used a questionnaire that had already demonstrated reliability. After collecting the questionnaires, the data was analyzed using STATA software version 14, and the results were presented in detail (see Table 2).

**Table 2:** Reliability Results.

| SN | Variable | Number of items | Cronbach's Alpha ($\alpha$) | Cronbach's Alpha ($\alpha$) on standardised items |
| --- | --- | --- | --- | --- |
| 1. | Data utilization to develop AHWPs | 13 | 0.90 | 0.93 |
| 2. | Health system related factors | 08 | 0.84 | 0.87 |

### Data management and analysis

The data management process for this study involved several steps to ensure the integrity, security, and organization of the data. Data was collected through the administration of questionnaires and a review of documents for evidence from respondents. It was then entered into an Excel dataset for storage and analysis. The data was reviewed for errors, inconsistencies, and missing values. Errors were corrected, and a few missing values were addressed through data imputation. Coding was performed using numerical values in STATA version 14. Regular backups were made on the computer to prevent data loss in case of technical issues or errors.

Under univariate analysis, descriptive statistics, including frequencies and percentages, were used to summarize the socio-demographic characteristics of the respondents. Data utilization for the development of Annual Health Work Plans (AHWPs) was also assessed using descriptive analysis and reported in frequencies and percentages. The analysis of results involved multiple stages to determine the health system-related factors influencing data utilization for AHWP development. The model-building process included both bivariate and multivariate analyses, as follows: Bivariate analysis was conducted to identify health system-related factors influencing data utilization for AHWP development. This analysis determined which variables had a statistically significant relationship with data utilization. Crude Odds Ratios (COR) were calculated for each variable to quantify the strength of the association between the independent variable and the outcome. A p-value was computed for each factor to assess statistical significance, with a threshold of p < 0.05. Additionally, 95% confidence intervals (CI) were reported to indicate the range within which the true association likely lies.

Multivariate analysis was then performed to control for confounding factors and identify variables that remained independently associated with data utilization after adjusting for other factors. This analysis utilized Adjusted Odds Ratios (AOR) to assess the impact of each predictor while accounting for the effects of other variables. Variables that were statistically significant at the bivariate level (p < 0.20) were included in the multivariate logistic regression model. This step was crucial in building a final model that explained the determinants of data utilization for AHWP development.

This structured approach to model building ensured that the final set of predictors provided a comprehensive understanding of the factors influencing data utilization. The use of both bivariate and multivariate analyses allowed the study to identify key drivers of data utilization while accounting for potential confounders.

### Ethical considerations

Ethical approval was obtained from the Uganda Christian University-Research Ethics Committee for clearance under UCUREC-2024-914 before the commencement of the study. Informed written consent was obtained from all study participants, and their confidentiality and privacy were safeguarded throughout the study. Participants were given the right to voluntary participation and if they wished to withdraw from the study any time without any penalty.

## Results

### Socio-demographic characteristics of respondents

The demographic profile of the 313 respondents (95.7%) reveals a predominantly Catholic population (46.65%), with most being married (77.96%) and female (59.74%). The majority (82.43%) reside outside health center quarters, and a significant proportion of health workers (91.69%) do not have medical records qualifications. Smaller proportions among respondents include Anglicans (37.70%), Muslims (15.65%), single individuals (22.04%), males (40.26%), and medical records cadres (8.31%). See Table 3.

**Table 3:** Socio-demographic characteristics of respondents.

| Variable | Category | Frequency | Percent (%) |
| --- | --- | --- | --- |
| n=313 |  |  |  |
| Religion | Anglican | 118 | 37.70 |
|  | Catholic | 146 | 46.65 |
|  | Muslim | 49 | 15.65 |
| Marital status | Married | 244 | 77.96 |
|  | Single | 69 | 22.04 |
| Residence | Quarters | 55 | 17.57 |
|  | Outside quarter | 258 | 82.43 |
| Gender | Males | 126 | 40.26 |
|  | Females | 187 | 59.74 |
| Cadres | Medical Records | 26 | 8.31 |
|  | Other health workers | 287 | 91.69 |

### The level of data utilization for developing health facility annual work plans

Examining the level of data utilization for AHWP development in health centers through univariate analysis revealed that 97 out of 313 respondents (30.99%) utilized data, while 69.01% did not. See Figure 1.

**Figure 1:**
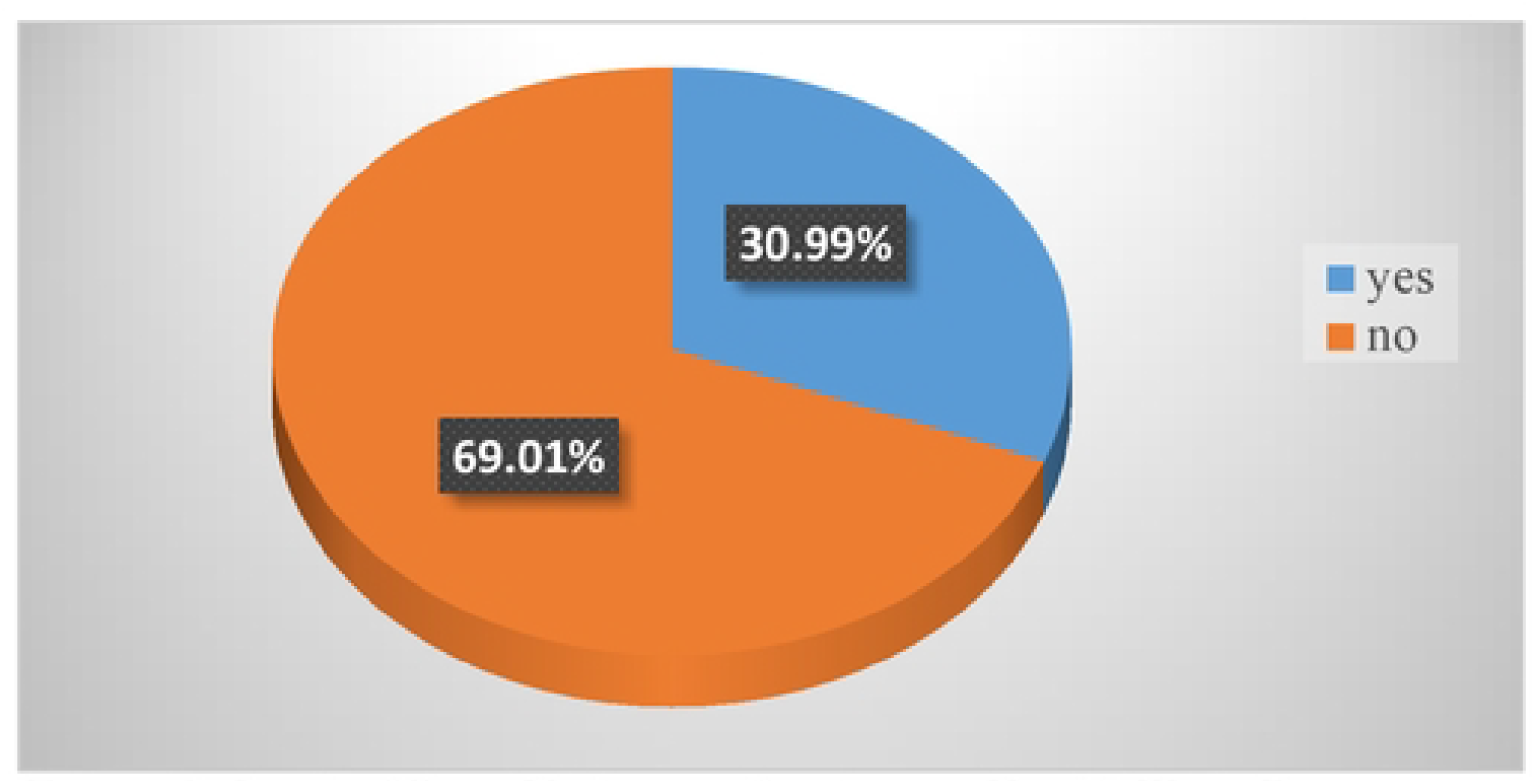
Level of Data Utilization for Annual Health Work Plans

### Health system-related factors influencing data utilization for AHWPs development

This section presents the study results, highlighting the health system-related factors that influence data utilization for Annual Health Work Plan (AHWP) development, as determined through bivariate and multivariate analysis. In the bivariate analysis, several health system-related factors were found to statistically influence the utilization of data for AHWP development in health centers in the Bukedi region. These factors included the frequency of facility meetings, data quality, feedback from the Ministry of Health (MOH), facility leadership, staff training in data utilization, and internet availability.

Regarding facility meetings, it was surprising to find that those who had regular meetings were 0.45 times less likely to utilize data (COR: 0.45; 95% CI: 0.22-0.91; P=0.028) compared to those who never had meetings. In terms of data quality, those with poor data quality were 0.0008 times less likely to utilize data (COR: 0.0008; 95% CI: 0.0002-0.004; P=0.000) compared to those who did not face poor data quality issues. Regarding feedback from the MOH, respondents who received feedback on data utilization were 40.8 times more likely to utilize data (COR: 40.8; 95% CI: 17.4-95.8; P=0.000) compared to those who never received such feedback. For leadership, facilities with leaders who fostered a culture of data utilization were 36.7 times more likely to utilize data (COR: 36.7; 95% CI: 18.6-72.4; P=0.000). Additionally, facilities with staff trained in data utilization were 265.5 times more likely to use data (COR: 265.5; 95% CI: 92.8-759; P=0.000), while facilities with internet availability were 0.06 times less likely to utilize data (COR: 0.06; 95% CI: 0.009-0.48; P=0.007) compared to those without internet access. See Table 4.

**Table 4:**
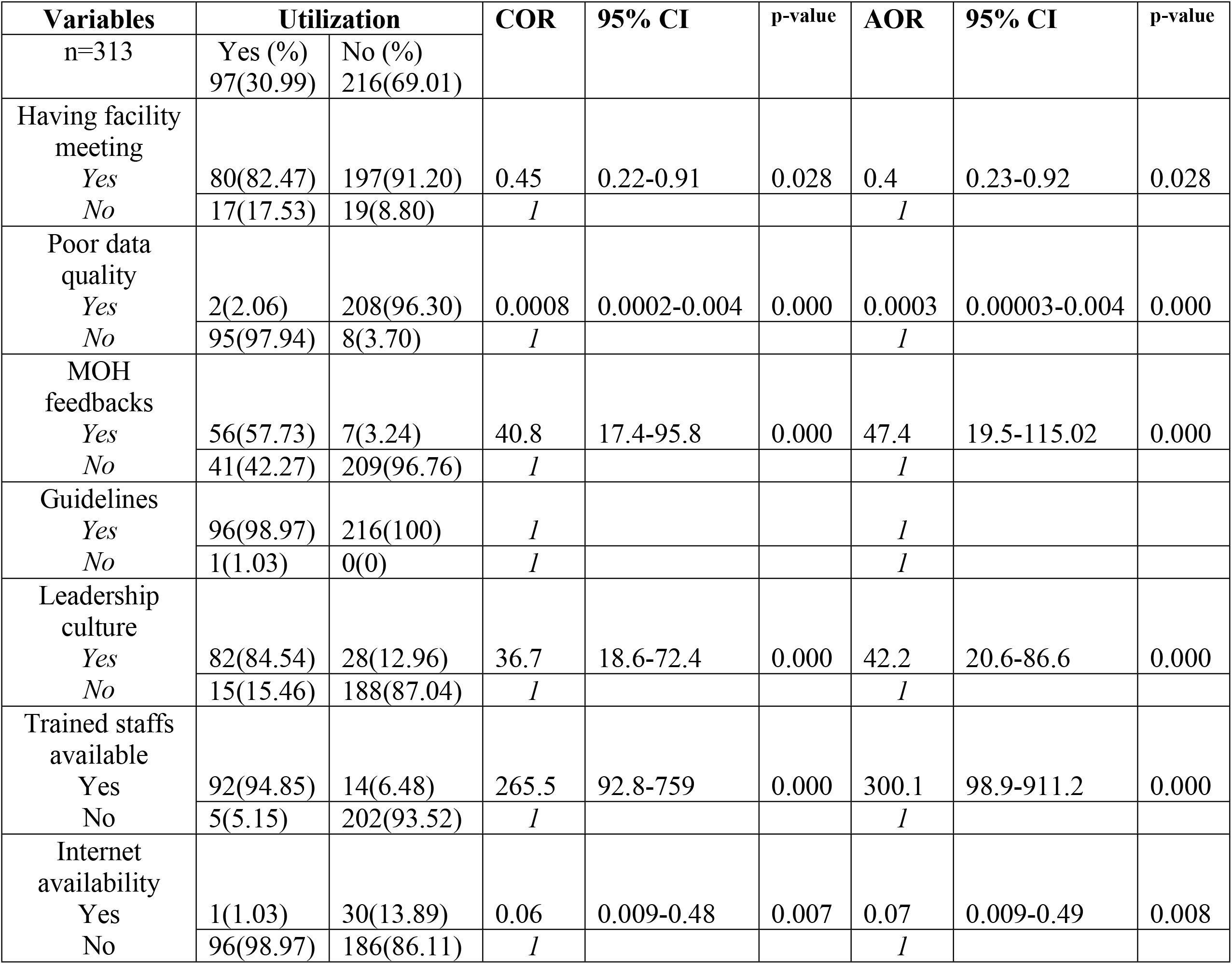
Bivariate and multivariate analysis of health system related factors influencing data utilization for AHWP development.

At multivariate analysis, several health system-related factors were statistically associated with the utilization of data for Annual Health Work Plan (AHWP) development in health centers in the Bukedi region. Facilities that held regular meetings were 0.4 times less likely to utilize data (AOR: 0.4; 95% CI: 0.23-0.92; P=0.028) compared to those that never had meetings. Similarly, facilities with poor data quality were 0.0003 times less likely to utilize data (AOR: 0.0003; 95% CI: 0.00003-0.004; P=0.000) compared to those that never experienced poor data quality issues. Interestingly, facilities that received feedback from the Ministry of Health (MOH) regarding data utilization were 47.4 times more likely to utilize data (AOR: 47.4; 95% CI: 19.5-115.02; P=0.000) compared to those that never received feedback. See Table 4.

Furthermore, facilities with a leadership culture of data utilization were 42.2 times more likely to use data (AOR: 42.2; 95% CI: 20.6-86.6; P=0.000) compared to those without a culture of data use. Facilities with staff trained in data utilization were 300.1 times more likely to utilize data (AOR: 300.1; 95% CI: 98.9-911.2; P=0.000) compared to those without trained staff. Lastly, facilities with internet access were 0.07 times less likely to utilize data (AOR: 0.07; 95% CI: 0.009-0.49; P=0.008) compared to those without internet access. See Table 4.

## Discussion

This study aimed to determine the level of utilization of routine health data and its associated factors among healthcare workers in health centers in the Bukedi region. The findings from Bukedi, where only 30.99% of participants utilized data to develop Annual Health Work Plans, reveal that data utilization falls short of the ministry of health planning tool HMIS001 and PHC sector guidelines, which recommend data utilization rates of 80% and above. This reflects a gap in the use of data for health planning and interventions. These results are consistent with broader trends observed in Uganda and other low- and middle-income countries, as highlighted in a study by Ayele and others, which reported data utilization rates ranging from 38.4% in Northern Ethiopia to 48.1% in Kenya [14].

The low data utilization rate (30.99%) for AHWPs development in Bukedi, Uganda, can be attributed to several factors, including ineffective leadership by managers and challenges related to poor-quality data in health centers, which may discourage healthcare workers from using data for health planning. Additionally, healthcare workers may lack the necessary skills to effectively utilize data for health planning, as observed in this study. However, the data utilization rate in Bukedi is lower than that observed in studies from the North Gondar Zone in the Amhara region (78.5%) and the Hadiya Zone (69.3%) [5, 15]. This discrepancy could be due to differences in the type of decisions the data was used for, the type of facility, the participants involved, and the criteria used to measure data utilization.

Importantly, it has been suggested that the Health Information System Strengthening Model (HISSM) strengthens health information systems by integrating diverse health data sources, including facility, community, electronic health records, population-based data, human resources, financial, supply chain, and surveillance data, to support informed decision-making. This highlights the lack of a similar approach in health centers in the Bukedi region. Perhaps, adopting such models in Bukedi could improve data utilization for AHWPs development.

Meetings influenced data utilization for AHWPs development; however, an unexpected result was observed: health facilities that conducted regular meetings had significantly lower odds of utilizing data for their annual health work plans compared to facilities that did not hold regular meetings. This suggests that simply having regular meetings does not automatically lead to effective data use in health planning. This could be due to the fact that the content of the facility meetings may not focus on data analysis or health planning, or the meetings may be predominantly administrative, addressing issues unrelated to data utilization. Such meetings may not contribute to the improved use of data. Furthermore, frequent meetings without clear objectives, agendas, or actionable outcomes may be less effective than fewer, well-organized meetings that prioritize data-driven decision-making. Notably, some regular meetings might create complacency among staff, who may feel that merely attending meetings fulfills their responsibilities, even if data is not being effectively analyzed or applied.

Contrary to the findings above, several studies have found that regular meetings positively influence the utilization of data for decision-making and health planning. Regular meetings provide a platform for staff to discuss data findings, share insights, and collaborate on interpreting results, as noted in Mphatswe et al.’s study, which observed that facilities conducting routine data review meetings saw improvements in data quality and utilization as staff collaboratively identified issues and developed solutions [16]. Similarly, regular meetings can serve as training opportunities where staff enhance their skills in data analysis and interpretation, as seen in Gimbel et al.’s study in Mozambique, which found that incorporating data discussions into regular meetings fostered a data-driven culture and improved staff competencies [17]. Regular data discussions in meetings promote accountability among health workers, leading to increased data utilization through feedback mechanisms and cultivating an organizational culture that values evidence-based practices, which are essential for sustained data use in health systems [18]. Although the study’s initial analysis suggests that regular meetings do not necessarily enhance data utilization for annual health work plan development, evidence from other studies indicates that meetings can positively influence data use when they are well-structured and focused on data-related activities. Therefore, healthcare staff in the Bukedi region should adopt this model for their meetings.

The study findings also revealed that, in health centers in the Bukedi region, facilities that experienced poor data quality—such as incomplete data or lack of access to reliable data—had significantly lower odds of utilizing data for annual health work plans compared to those without data quality issues. This result concurs with a study conducted in Eastern Ethiopia, which found that data quality, among other factors, significantly influenced the utilization of health data [19]. This stark difference highlights the profound impact that data quality has on the confidence and ability of health workers to use data effectively. This can likely be attributed to various factors, such as inaccurate and unreliable data, which can lead to misleading or faulty information. This makes it challenging for health workers to make informed decisions, as incomplete and inconsistent data can create confusion and mistrust, reducing the willingness and ability to use it to identify trends, patterns, and poorly performing indicators needed for data-driven decision-making required in developing annual health work plans.

This is supported by Ndabarora and others in their study in LMICs, who noted that errors and inconsistencies in the data can cause health workers to distrust the information, leading them to avoid using it in decision-making processes [20]. Similarly, untimely and outdated data can result in delayed decision-making, reducing its relevance and making it difficult for health workers to use data for planning and decision-making embedded in the annual health facility work plans. This finding supports the literature, which highlights the importance of data quality in promoting data utilization, as demonstrated by Nutley and Reynolds, and Asiimwe in Uganda [18, 21]. The statistical significance of poor data quality suggests that this factor is a critical predictor of data utilization, and addressing it could positively impact health facility planning and decision-making for the allocation of resources and inputs, ultimately enhancing service delivery.

Contrary to the findings above, some studies suggest that poor data quality does not always lead to low data utilization. Scott et al. observed that, in some contexts, health professionals continue to use available data for decision-making despite quality issues, often supplementing it with local knowledge and experience [22]. Additionally, Lippeveld argued that the relationship between data quality and utilization is not always linear and requires complementary efforts to promote data demand and create an information-driven culture to realize the benefits of improved data quality [23]. This could possibly be the missing link among health workers in the Bukedi region, who may rely on experience and knowledge, alongside other measures, to enhance data utilization for AHWPs development in their health centers.

The multivariate analysis further indicates that participants from health facilities that received feedback from the Ministry of Health about data utilization had significantly higher odds of using data for annual health work plans compared to those that did not receive such feedback. This finding aligns with Mekonnen’s and Gebeyehu’s systematic review and meta-analysis, which identified regular top-level feedback as a key factor for enhanced utilization of routine health information [24]. This could be attributed to the positive impact of feedback from top-level technical officers, which fosters accountability, improves skills, enhances communication, identifies data quality issues, and motivates health workers. In the end, this leads to more effective data-driven decision-making among health workers. This is supported by various studies, including one by Hotchkiss et al., which demonstrated that feedback from the Ministry of Health creates a sense of accountability among health workers, encouraging them to use data more effectively. The knowledge that their data contributions are reviewed and valued at higher levels motivates staff to prioritize data use [25]. Similarly, top-level feedback often includes recommendations and guidance on data management, usually presented in debrief meetings and documented in support supervision books. This can improve health workers’ skills and competencies in data analysis and interpretation. Furthermore, Lippeveld observed that feedback can help facilities identify errors and inconsistencies in their data, allowing them to address these issues promptly and improve overall data quality. Over time, this boosts morale and motivates health workers to continue using data for developing AHWPs [18, 23].

However, despite the general trend, some studies have found that feedback from higher authorities does not always lead to increased data utilization. It has been reported in some studies that top level authority feedback may be ineffective due to infrequency, lack of actionability and failing to address local needs, but also neglect local facilities situations, limiting impact of feedback [26]. Furthermore, excessive or overly critical feedback can demotivate health workers, while negative feedback without guidance is worthless [27]. These contrasting findings suggest that the effectiveness of feedback from higher authorities depends on its quality, relevance, and the way it is communicated. Feedback that is timely, specific, and supportive is more likely to enhance data utilization than feedback that is delayed or punitive.

The findings highlight a central role of MOH feedback in enhancing the utilization of health data for decision-making and planning. Feedback can boost data utilization, but effectiveness requires regular, specific, and constructive feedback. Health centres should develop tailored feedback processes to foster evidence-based decision-making.

Study findings further revealed a significant association between the presence of a leadership culture that emphasizes data utilization and the effective use of data in annual health work plans. In particular, participants who work in health facilities that fostered a leadership culture of data use had substantially higher odds of utilizing data compared to those that did not cultivate such a culture. This finding is consistent with existing literature by Hoxha and others in their study conducted on understanding the challenges associated with the use of health data in low- and middle-income countries which found which found that organizational culture, leadership, and resources played a role in influencing data utilization [28]. This could be probably because health facilities led by leaders who prioritize data use are able to create a supportive environment by prioritizing data use, allocating resources, providing training, promoting open communication, setting goals, leading by example, and fostering teamwork which in the end influences data utilization for developing AHWPs. In Gimbel and others, effective leaders allocate necessary resources, including training, technology, and infrastructure, to support data collection, management, and analysis [17]. Additionally, effective leadership cultivates a culture of accountability and motivation, driving health workers to prioritize data use. By setting clear expectations and recognizing data-driven achievements, leaders encourage staff to value data quality and utilization [23]. Moreover, leaders who champion data use foster a supportive environment, empowering staff to access, analyze, and apply data, promoting collaboration and evidence-based decision-making [25].

These trends supported by various studies as in Gimbel and others in Mozambique who reported that health facilities with proactive leadership were more likely to utilize data effectively for planning and resource allocation further observing that leadership support was instrumental in overcoming barriers to data use, such as limited technical skills and inadequate infrastructure [17]. Similarly, Hotchkiss and others demonstrated that leadership engagement is vital for the continuous improvement of health information systems where leaders who actively participate in data review meetings and decision-making processes set a precedent for their teams to follow suit [25].

Contrarily, while the majority of studies support the positive influence of leadership culture on data utilization, some research presents contrasting findings. It has been reported that, effective health facility leadership alone is insufficient to promote data utilization in health facilities, as entrenched organizational structures, resistance to change, superficial commitment, and top-down approaches can undermine efforts, highlighting the need for collaborative and sustained leadership engagement [27, 26, 29]. These contrasting findings indicate that leadership culture significantly promotes data utilization, but its success depends on authentic, consistent, and comprehensive leadership efforts, supported by systemic changes, genuine engagement, and a holistic approach to addressing barriers.

Consistent with existing literature, strong and committed leadership fosters an environment that prioritizes data use, allocates necessary resources, builds capacity, and holds staff accountable for data-driven outcomes. However, the effectiveness of leadership in promoting data utilization for developing AHWPs is not guaranteed and depends on the depth of commitment, the presence of supportive systems, and the ability to address some of the health system challenges. Health systems aiming to improve data utilization should focus on promoting a leadership culture that genuinely values and integrates data use into all aspects of health service delivery including development of AHWPs.

A significant association between staff training and data utilization for AHWPs development was also observed. Participants who reported having adequately trained staff in their facilities had substantially higher odds of utilizing data for annual health work plans compared to those without trained staff. This finding concurs with the findings reported by Asemahagn and Alene in Ethiopia which observed that availability of resources was significantly associated with the utilization of health data [19]. This could be probably because adequately trained staff in health facilities are more likely to utilize data effectively for AHWPs development due to teamwork, shared data utilization responsibility and accurate planning.

The findings are further supported by various studies that observed staff training significantly enhances data utilization by equipping health workers with essential skills and competencies for accurate data collection, management, analysis, and interpretation [27]. Additionally, training not only ensures high-quality data for effective health planning and resource allocation but also boosts staff morale and motivation, fosters a culture of evidence-based decision-making, and builds capacity for proactive problem-solving through data analysis [30, 31, 25].

While staff training is positively correlated with data utilization, research shows that training alone may not be sufficient to increase health data use. Studies have identified various barriers that hinder the effective application of training, including systemic and organizational issues. For example, high workloads, lack of management support, inadequate infrastructure, insufficient incentives, and entrenched decision-making processes can limit data utilization despite training [32, 33]. These findings suggest the need for comprehensive approaches that address underlying organizational barriers and support sustained changes in data-driven practices.

Interestingly, the study findings also revealed that participants from health facilities with internet access had significantly lower odds of utilizing data for annual health work plans compared to those without internet access. This could be due to several factors, including the possibility that some health workers lacked computer skills, had no training on data systems, or resisted new technology. Additionally, poor internet connectivity and inadequate equipment may have further complicated data use for AHWPs development. Healthcare data utilization is hindered by several key obstacles, such as limited technical skills among health workers, insufficient training on digital health systems, resistance to change due to familiarity or technological fears, and infrastructure challenges like unreliable electricity and slow internet. These interconnected barriers must be addressed to enhance healthcare data utilization and improve patient outcomes [22, 31, 26, 34].

This unexpected finding is supported by previous studies, which indicate that merely having internet access does not ensure efficient data use. Kimaro and Sahay found that in Tanzania, the implementation of internet-enabled health information systems did not significantly improve data use due to organizational and socio-cultural barriers [26]. Despite having internet access, health workers continued to rely on traditional paper-based methods. Similarly, Chib et al. observed that internet connectivity did not enhance data utilization in rural health facilities in Asia because health workers lacked the necessary training and support to use digital tools effectively.

Conversely, studies in Mozambique, Ethiopia, and elsewhere indicate that internet availability can enhance data utilization, but its effectiveness depends on complementary factors such as training, system design, and organizational culture, highlighting variability in outcomes [17, 35, 31].

In conclusion, internet access alone is not sufficient for effective healthcare data utilization for AHWPs development in health centers in the Bukedi region. A holistic approach is needed, including comprehensive training, user-friendly systems, reliable infrastructure, change management, leadership support, and cultural shifts. These elements are essential to leveraging internet availability and enhancing data-driven decision-making for improved health planning.

## Conclusions

The findings revealed a significant gap in data utilization, with only 30.99% of healthcare workers using routine data for AHWP development—far below the desired utilization rate of 80%. This low utilization rate can be attributed to factors such as insufficient training, poor data quality, ineffective leadership, inadequate infrastructure, and the absence of feedback mechanisms. The study highlights that effective leadership, supportive feedback, comprehensive training, and a culture that values data-driven decision-making are key determinants of data utilization for health planning. Additionally, infrastructure challenges, including limited internet connectivity and poor data management systems, remain significant barriers. To enhance data utilization in health planning, a holistic approach is required. Providing internet access or training programs in isolation is insufficient. Instead, leadership engagement, cultural shifts, adequate infrastructure, and supportive organizational environments must work together to foster a culture of data-driven decision-making. The adoption of models such as the Health Information System Strengthening Model (HISSM) can be instrumental in addressing these challenges

### Recommendations

1. Leadership and Supportive Supervision: Health facility leadership must prioritize data utilization by actively promoting and facilitating data-driven practices. Facility managers should receive training in leadership strategies that foster accountability and encourage data use through regular supervision and the establishment of clear data-related objectives.
2. Organizational Culture Change: Health facility leadership should cultivate a culture that values data utilization. Facility managers should lead by example, integrating data use into decision-making and performance evaluations. Highlighting success stories of data-driven healthcare improvements can further motivate staff to embrace data utilization.
3. Addressing Data Quality Issues: Efforts should be made at the facility level to enhance the quality of routine health data. This includes implementing standardized data collection procedures, conducting regular data audits, and ensuring data completeness, accuracy, and timeliness.
4. Capacity Building and Training Programs: District health teams should provide regular and comprehensive training to health workers to improve their skills in data analysis, management, and interpretation. Training programs should also focus on practical applications of data to demonstrate its relevance in health planning.
5. Feedback Mechanisms: District health teams should collaborate with the Ministry of Health to establish and maintain a robust feedback system for health facilities. Timely and constructive feedback on data collection and usage will help health workers identify gaps, make necessary improvements, and encourage greater use of health information.
6. Infrastructure Improvement: The Ministry of Health, in partnership with other stakeholders, should invest in enhancing infrastructure, including reliable internet connectivity and functional health information systems. Ensuring that healthcare workers have access to the necessary tools will facilitate effective data collection, access, and utilization.

### Suggested areas for future research

1. Barriers to Data Utilization: Further research is needed to explore the specific barriers at various levels of the health system that hinder data utilization among health workers. Identifying these barriers could help develop targeted interventions to improve data use for health planning.
2. The Role of Organizational Culture and Leadership: Investigating how organizational culture, leadership, and supportive work environments influence the use of routine health data for decision-making can help in designing more effective leadership and cultural change interventions to enhance data utilization.
3. Effectiveness of Feedback Mechanisms: Assessing the impact of feedback mechanisms from the Ministry of Health to health facilities in fostering data-driven decision-making could help determine best practices for providing actionable and motivating feedback.
4. Impact of Infrastructure and Technology: Research should examine how infrastructure and technology, such as internet access and electronic health information systems, influence data utilization for health planning. This could inform the design of more effective infrastructure support programs.
5. Long-term Impact of Training and Capacity-Building: Further research is needed to evaluate the long-term effects of training and capacity-building initiatives on health workers’ ability to utilize data effectively. Additionally, studies should focus on identifying the optimal content and frequency of training programs that lead to sustained data use in health planning.

## Data Availability

Data set will be accessed without restrictions.

## Abbreviations

AHWP: Annual Health Work Plan
HMIS: Health Management Information System
MOH: Ministry of Health
UHC: Universal Health Coverage
HIS: Health Information System
WHO: World Health Organisation
SDG: Sustainable Development Goal
HCs: Health Centres
HIQA: Health Information and Quality Authority
LMICs: Low and Middle Income Countries
HSTP: Health Sector and Transformation Plan
PHC: Primary Health Care
ANC: Antenatal Care
PNC: Postnatal Care
FP: Family Planning
COR: Crude Odds Ratio
AOR: Adjusted Odd Ratio
HISSM: Health Information system Strengthening Model.

## Acknowledgements

The authors extend their sincere appreciation to the data collectors, District Health Officers, Biostatisticians, In-charges of health centres and the rest health workers in Butebo and Pallisa districts for their support in data collection.

## Authors’ contributions

JC conceived the study idea. JC, NR, KMM, and SZ designed the study and wrote the protocol. JC, KMM, and SZ designed the data collection tools. JC and BA participated in data collection. JC, KMM, ZN, HMK, AW and BA undertook the analysis. JC, NR, BA, KMM, SN and ZN wrote the manuscript.

